# Pandemic-Driven Shifts in RSV Diversity, Evolution, and Global Spread

**DOI:** 10.64898/2026.11.02.26367099

**Authors:** Emanuele Gustani-Buss, Daniel M Weinberger, Nathan D Grubaugh, Vittoria Colizza, Yucai Shao, Marc Suchard, Anouk Evers, Natalie I. Mazur, Joanne G. Wildenbeest, Robert J. Lebbink, Hanneke J.A.A. van Zoggel, Louis Bont, INFORM-RSV Study Group, Philippe Lemey

## Abstract

The COVID-19 pandemic dramatically disrupted the seasonal circulation of RSV, with subsequent heterogeneous global resurgence. We aimed to characterize how pandemic-era disruptions restructured global RSV transmission dynamics, evolutionary patterns, and drivers of reemergence. We show that RSV-A and RSV-B genetic diversity underwent profound pandemic-associated suppression followed by heterogeneous resurgence. The post-pandemic period was characterized by pronounced changes in diversity, with the emergence of novel RSV-A genotypes (A.D.3 and A.D.5.2) and the expansion of the RSV-B, B.D.E.1 genotype. The recovery of RSV-B deviated strongly from neutral expectations and positive selection was detected at 20 sites in the G glycoprotein, F fusion protein, and M2-2 regulatory protein. Phylogeographic reconstruction revealed a sharp decline in international viral spread during the pandemic, followed by increasing persistence. Europe consistently served as the predominant global source, followed by Western Pacific and North America. Air-travel volume was the strongest predictor of dissemination (log coefficient >0.68 for both subtypes). Post-pandemic RSV resurgence represented fundamental epidemiological restructuring shaped by a population bottleneck, positive selection and major impact of air-travel. These findings underscore the need for robust genomic surveillance to track the global circulation of lineages, and to monitor effectiveness of pharmaceutical interventions given the emergence of new genotypes.

**One Sentence Summary:** Air Mobility and Diversifying Selection Shape Post-Pandemic RSV Global Diversity

## Introduction

Human respiratory syncytial virus (RSV) is a major viral pathogen associated with acute lower respiratory infections, imposing a substantial global disease burden, particularly among young children under 5 years and adults over 65 years. RSV epidemics typically peak during the autumn-winter season from April to June in temperate regions of the Southern Hemisphere and from October to February in the Northern Hemisphere ^1,2^. Human behavior represents a critical driver of RSV transmission, including mobility, indoor aggregation, and population density, alongside environmental conditions such as temperature and humidity ^1,3^. Previous work empirically demonstrated that air travel mobility plays a crucial role in RSV global dissemination patterns ^4^.

Respiratory virus transmission, including RSV, was significantly disrupted by the COVID-19 pandemic in 2020. Unprecedented changes in human behavior resulted from non-pharmaceutical interventions (NPIs), such as lockdowns, school closures, social distancing, and international travel restrictions, which constrained transmission of SARS-CoV-2 and other respiratory viruses. For influenza virus, global positivity rates were reduced by approximately 95% during early pandemic phase, representing the most dramatic suppression of influenza activity ever recorded in the modern surveillance era ^5^. Yet critically, the COVID-19 pandemic did not merely suppress influenza virus but fundamentally restructured its global transmission architecture. Phylogeographic analyses revealed a marked contraction in the diversity of globally circulating clades, a near-complete interruption of inter-regional lineage exchange, a profound disruption of established source-sink transmission dynamics, and a collapse of lineage persistence across successive seasons ^5^.

Following the relaxation of pandemic-related measures, RSV resurged heterogeneously across countries, yet the full consequences of this disruption on global RSV dynamics remain poorly understood. The selective pressures governing viral adaptation during and after the population bottleneck, and their implications for RSV subtype fitness, have yet to be elucidated. We hypothesize that pandemic-induced changes in human behavior and mobility shifted RSV lineage circulation dynamics. To address these gaps, we present a comprehensive multicenter genome-wide analysis of approximately 1,320 RSV isolates spanning eighteen countries from 2020 to 2023, representing the most geographically comprehensive RSV complete genome dataset assembled to date. By integrating complete genome sequences with international air travel data within a Bayesian phylodynamic framework, we characterize spatiotemporal population structure, lineage persistence, evolutionary rate variation, and selection pressures across four epidemiologically distinct periods. Our analyses reveal distinct evolutionary trajectories between RSV-A and RSV-B, with positive selective pressure in key functional genes potentially underlying the competitive dominance of RSV-B observed post-pandemic.

## Results

### RSV epidemiological landscape

RSV case numbers illustrate marked seasonal trends in RSV positivity across the WHO regions prior to 2020, in line with typical RSV seasonality (Fig. 1A, FluNet data through the Global Influenza Surveillance and Response System (GISRS)). In 2020, RSV cases declined sharply, coinciding with the COVID-19 pandemic and the implementation of non-pharmaceutical interventions (NPIs), such as face-masking, social distancing and travel bans. These measures caused an unprecedented disruption in RSV transmission. From 2021, RSV cases re-emerged with variable intensity across years and regions, suggesting heterogeneous, asynchronous and altered seasonal patterns (Fig. 1A).

**Fig. 1.**
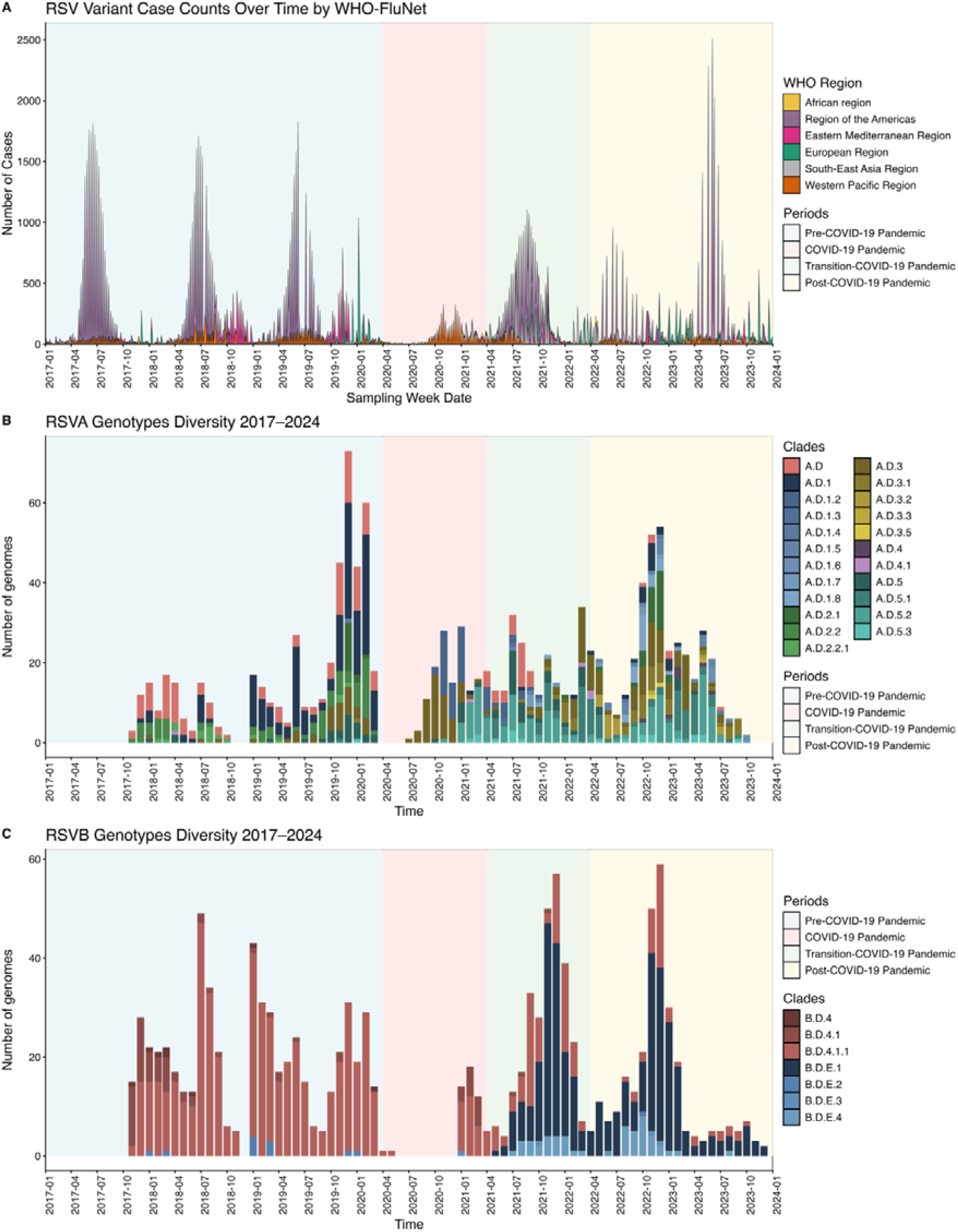
Global respiratory syncytial virus RSV genomic epidemiology and case distribution, 2017-2024. A. Temporal trends in RSV cases reported through WHO’s FluNet surveillance system across six geographic regions. B. Temporal distribution of RSV-A genotypes from the INFORM surveillance study. C. Temporal distribution of RSV-B genotypes from the INFORM surveillance study.

To investigate how the COVID-19 pandemic and human behavior reshaped global RSV circulation dynamics, we compiled multicenter genomic datasets based on the INFORM study and publicly available genomes. Our datasets comprised genomes sampled from January 2017 to January 2024 and therefore covered different periods which we labeled as pre-pandemic (2017-2020), pandemic (2020-2021), transition (2021-2022), and relaxation/post-pandemic (2022-2024).

Similar fluctuating patterns were reproduced when summarizing the genomic sampling over time and its genotype assignment using Nextclade ^6^. The frequency distribution of RSV-A genotypes over time indicated a relative stability between 2017 and 2019 (Fig. 1B). This was followed by a marked drop in early 2020, with an increase in frequency in late 2020. The transition and post-pandemic periods (2021–2023) coincided with the re-emergence of multiple RSV-A lineages following a period of suppression during the pandemic. New genotypes (e.g., A.D.1.4, A.D.3.5, A.D.5.2, A.D.5.3) also emerged during the pandemic periods and intensified circulation in the transition to the post-pandemic period. Analysis of the 709 RSV-A genomes (2020–2023) included in this study revealed a total of 23 distinct sub-lineages. Among RSV-A sub-lineages, A.D.3 was the most frequently detected (22.14%, n=157/709), persisting throughout all study periods, followed by A.D.5.2 (19.75%, n=140/709) (Table S1). Compared to RSV-A, RSV-B shows a lower degree of genotype diversification (n = 8) based on 611 genomes, largely dominated by B.D.E.1 in (59.57%, n=364/611) and B.D.4.1.1 (27.66%, n=169/611) (Fig. 1C, Table S2). Periodic fluctuations in RSV-B genome numbers are visible, with a pronounced drop during 2020 and a resurgence thereafter. The emergence of additional subclades (e.g., B.D.E.1, B.D.E.3, B.D.E.4) post-pandemic suggests that RSV-B, like RSV-A, was undergoing diversification upon resurgence of transmission.

### RSV phylodynamics revealed persisting genotypes in the post-pandemic period

The RSV-A maximum credibility tree showed substantial diversity across multiple co-circulating clades (A.D.1, A.D.3 and A.D.5.2) throughout the 2017-2024 period (Fig. 2A). Although some clades demonstrated regional clustering, most notably those circulating in North America, South America, and Europe, the overall phylogeographic pattern was characterized by geographically dispersed lineages across most clades. The transition period (2021-2022) was marked by clades comprising lineages persisting through the period of stringent non-pharmaceutical interventions. The post-pandemic period (2022–2024) was characterised by phylogenetic expansion across all major clades, with renewed geographic mixing within transmission clusters. Notably, several clades exhibited intercontinental spread, with samples from multiple regions, particularly for A.D.5.2, which showed frequent long-distance dispersal events.

**Fig. 2.**
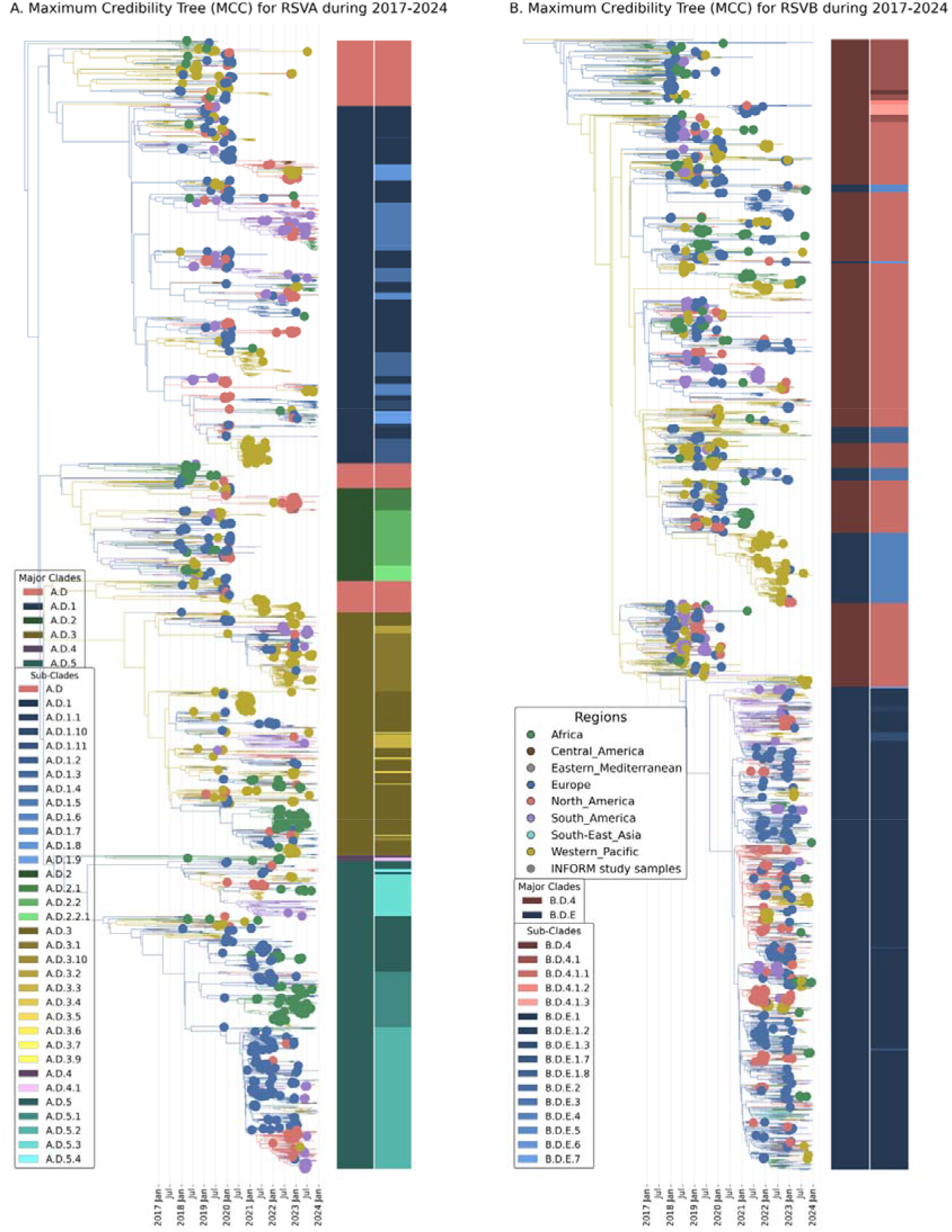
Maximum Credibility Trees (MCC) for RSV A and B during 2017-2024. Phylogenetic reconstruction of (A) RSV-A and (B) RSV-B evolution from 2017-2024 using Bayesian evolutionary analysis. Tips represent viral sequences colored by geographic region. Tip points are sized to highlight sequences from the INFORM study (larger points). Colored bars adjacent to tips indicate genetic clade assignments. The trees are time-scaled with x-axis showing sampling dates from January 2017 to January 2024. Branch colors correspond to the most probable geographic region state at each node, reconstructed using discrete trait analysis. The phylogenies illustrate the temporal distribution, geographic spread, and clade dynamics of RSV circulation before, during, and after the pandemic period.

Like RSV-A, RSV-B underwent a clear evolutionary bottleneck during the pandemic period. However, the resurgence patterns differed markedly between subtypes; while RSV-A recovery was driven by multiple surviving lineages, RSV-B resurgence was predominantly attributed to expansion of a single major lineage (Fig. 2B). The phylogeny revealed a large well-supported monophyletic clade (B.D.E.1, posterior probability = 1.0) that emerged in early 2020 and became globally predominant during 2021-2023, comprising 1535 genomes collected worldwide during the transition and post-COVID-19 pandemic (2021–2023, among them 364 samples for our study). This clade exhibited extensive geographic mixing, with samples from Africa, Europe, North America, South America, and Asia interspersed throughout, suggesting rapid international dissemination following emergence.

Lineage diversity, however, was not entirely lost during the pandemic period. Several phylogenetically distinct clades (B.D.4, B.D.4.1.1, B.D.4.1.2, B.D.4.1.3, B.D.E.3, B.D.E.4, B.D.E.5, B.D.E.6, B.D.E.7) persisted at lower frequencies and co-circulated with the dominant B.D.E.1 lineage during the post-pandemic period. These persistent lineages demonstrated variable geographic distributions, with some maintaining region-specific circulation patterns while others achieved broader geographic spread. The phylogenetic structure suggests that while B.D.E.1 benefited from a selective or stochastic advantage during the pandemic bottleneck, pre-existing lineage diversity was maintained at low levels and re-emerged as transmission intensity increased.

### Positive selection driving adaptation in the new genotypes in both RSV-A and RSV-B after the pandemic period

To elucidate potential molecular adaptation, we applied FUBAR and MEME selection detection methods to identify amino acid sites under positive selection across recent RSV-A RSV-B clades, focusing on sites supported by both analytical approaches. Our analysis focused on clades circulating after the pandemic period (Table S3). Clade A.D.5.2 showed three sites with positive selection supported by both methods (273, 297) in the G protein and one site in F protein (107). The A.D.3 clade demonstrated more extensive positive selection, particularly in the G protein with sites 132, 233, 273, 310, 313, and 314. The M2-1 protein in this clade also exhibited selection at position 128. Clade A.D.3.1 showed four G protein sites under selection (273, 284, 297, 298), while the SH protein displayed selection at position 55. In A.D.1.5, we only identified one in G (284) and one in L (1524). Clade A.D.1.3 showed more focused selection with site 271 in the G protein (with MEME identifying the closely related site 281). The position 273 at G protein was detected across multiple clades (A.D.5.2, A.D.3, A.D.3.1). RSV-B showed a different pattern for B.D.E.1 with 20 selected sites distributed across multiple proteins, including 10 in the G protein concentrated in positions 114, 145, 202, 217, 249, 256, 265, 275, 281 and one in F (42). Unlike RSV-A, RSV-B showed positive selection in replication machinery M2-1 (175) and M2-2 (48).

### The post-COVID pandemic RSV resurgence is shaped by selection

To assess whether RSV resurgence post-COVID-19 pandemic was shaped by non-neutral evolutionary dynamics, we applied a posterior predictive simulation procedure. This procedure used an entropy measure to summarize the distribution of descendants of lineages that were circulating during COVID-19 pandemic. We compared the entropy measure for our reconstructions to the neutral expectations under the same population size trajectories. While entropies for both RSV-A and RSV-B deviated from neutrality (Figure 3), we estimate a particularly strong impact of selection in the resurgence of RSV-B post-pandemic. This indicates that the emergence of the dominant RSV-B clade B.D.E.1 did not follow neutral expectations.

**Fig. 3.**
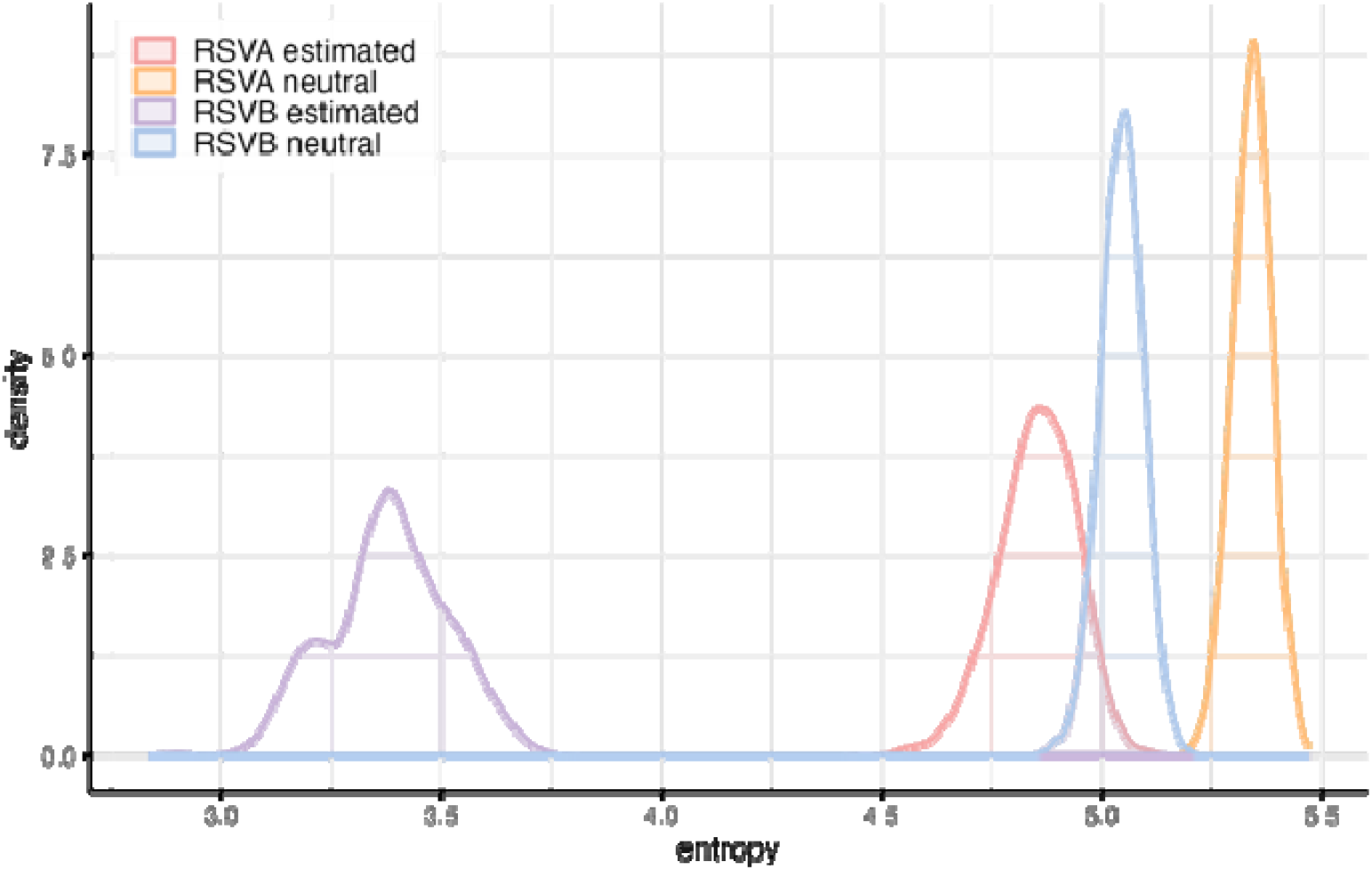
Assessment of deviation from neutral evolutionary expectations for RSV-A and RSV-B during the post-pandemic resurgence period. A Bayesian posterior predictive framework was used to disentangle the effects of population demographic history from genuine signatures of natural selection, integrating epidemic processes, evolutionary dynamics, and selective pressures through phylogenetic tree topology.

This pattern is consistent with the emergence and dominance of B.D.E.1, which suggests that key adaptive mutations arose prior to or early in the clade’s formation, enabling this lineage to successfully navigate the pandemic bottleneck. Subsequently, the positive selection observed within the established B.D.E.1 clade represents a distinct evolutionary phase, driving further adaptation and lineage diversification in the post-bottleneck environment.

### Global demographic dynamics

The demographic reconstructions show relatively consistent year-round circulation with some elevation in effective population size during certain periods for our 3,544 sequences for RSV-A and 3,800 genomes for RSV-B, covering eight global regions following the WHO designation: Africa, Central America, Eastern Mediterranean, Europe, North America, South America, South-East Asia and Western Pacific (Fig. 4A). This pattern likely reflects the aggregation of opposing seasonal dynamics from both hemispheres, resulting in temporally dispersed epidemic peaks that sustain continuous global transmission (Fig. 4B and C). The effective population size of RSV-A and RSV-B displayed remarkably similar temporal patterns within each geographic region, with overlapping median estimates and credible intervals in pre-pandemic period (Fig. 4A). In the pandemic period, there is a marked drop in population size around mid-2020 for both RSV-A and RSV-B, coinciding temporally with the onset of the COVID-19 pandemic. This period was characterized by the implementation of non-pharmaceutical interventions (e.g., lockdowns, school closures, and social distancing). A sharp rebound in effective population size began in late 2020, indicating resurgent transmission following the relaxation of COVID-19 control measures (Fig. 4A).

**Fig. 4.**
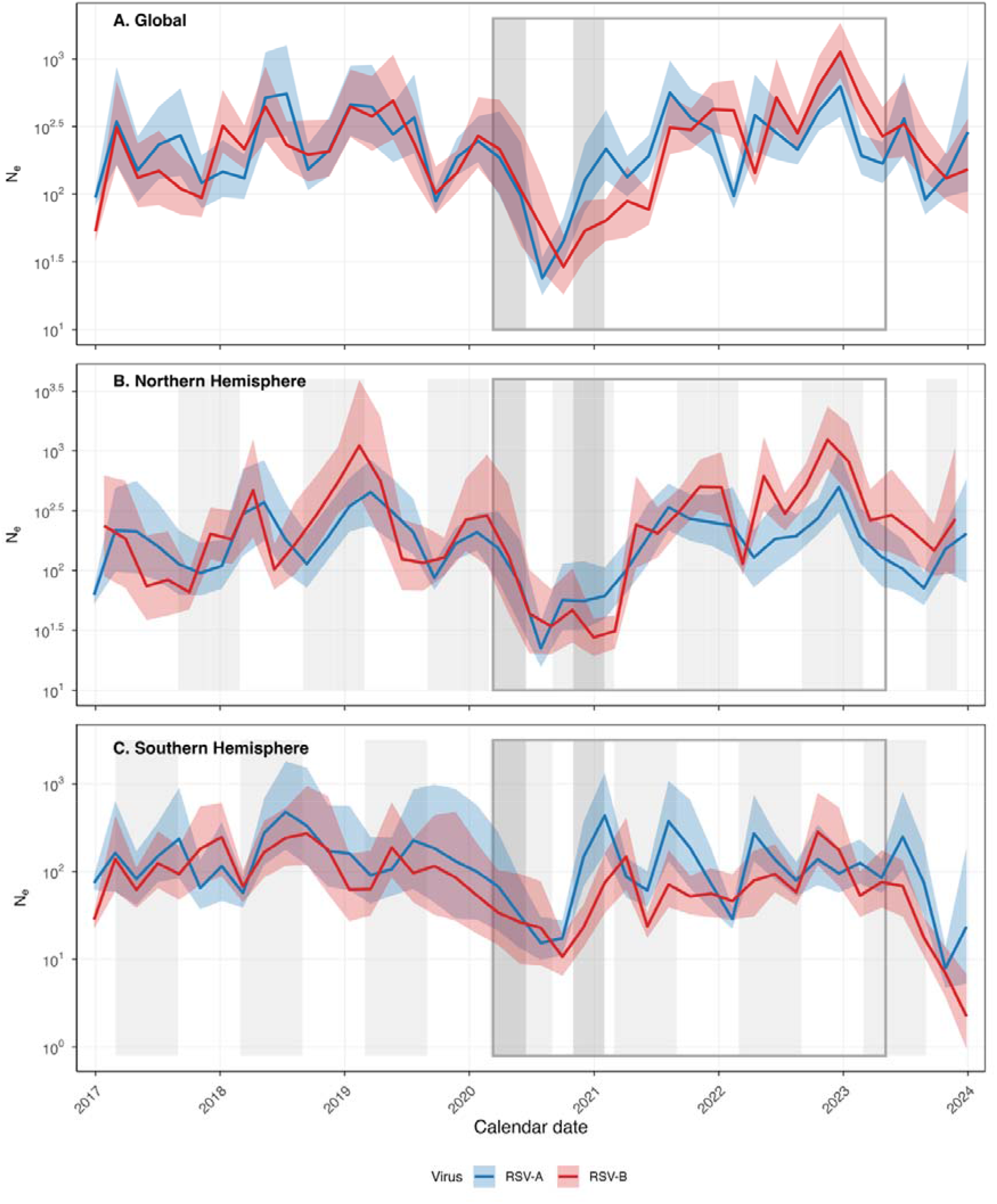
Effective population size (Ne) estimates through time under the Skygrid model for RSV-A and RSV-B across different geographical regions. (A)Global, (B) Northern Hemisphere, and (C) Southern Hemisphere. Notable reductions in effective population size during 2020-2021 correspond to non-pharmaceutical interventions implemented during the COVID-19 pandemic, followed by subsequent resurgence as restrictions were lifted. Lines represent the posterior mean of log-transformed effective population size (log(Ne)) over time for RSV-A (blue) and RSV-B (red). Shaded ribbons around each line indicate the 95% highest posterior density (HPD) intervals. Light grey vertical bands denote autumn-winter seasons (September–February) in the Northern Hemisphere. The dark grey outlined rectangle marks the WHO-defined COVID-19 pandemic period (March 11, 2020 – May 5, 2023). Dark grey filled rectangles indicate major lockdown periods (March–June 2020 and November 2020–January 2021).

Notably, the rebound for RSV-B appears to achieve higher population sizes, with high effective population size values continuing through 2022 and 2023 in the Northern Hemisphere (Fig. 4A). RSV-A also shows a strong resurgence with marked transmission dynamics in Southern Hemisphere (Fig. 4C).

### Spatial global dynamics-Unveiling the main routes of RSV transmission upon rebound

We implemented a Bayesian phylogeographic discrete approach to characterize the global population dynamics of RSV. Estimates of inter-regional migration events per unit time were derived from posterior summaries of transitions (Markov Jumps) for four periods: pre-pandemic, acute pandemic, pandemic transition, and post-pandemic periods. The discrete migration estimates underscored a relatively high number of viral movements pre-pandemic for RSV-A with multiple transitions from Europe (128 events, 95% Highest Posterior Density (HPD) interval = [80–130]) in our sample, followed by the Western- Pacific region (76 events, 95% HPD interval = [57–100]) (Fig. 5). The pandemic period indicates a marked decline in global transmission, with Europe, accounting for 24 events (95% HPD interval = [8–38]) and other regions contributing 1-2 introductions each (Fig. 5B). In the Transition period (2021), viral transmission appeared to increase slightly. Transitions from Europe doubled compared to estimates for the previous period (45 events, 95% HPD interval = [29-64]). While South America (9 events, 95% HPD interval = [3-17]), North America, and the Western Pacific region each accounted for 6 to 9 transmission events, as inferred sources of international RSV dissemination, representing a comparatively smaller contribution relative to the leading source regions (Fig. 5.C). The post-pandemic period (2022-2024) was characterized by an increase in transmission originating from Europe, South America, and North America. Europe remained the dominant source of global exportation events, with 52 events (95% HPD interval = [20-62]), followed by North America (33 events, 95% HPD interval 17-36), Western Pacific (24 events, 95% HPD interval 11-43), and South America (28 events, 95% HPD interval 10-36) (Fig. 5.D). Africa, Central America, and the Eastern Mediterranean exhibited minimal seeding activity, which likely reflects sampling limitations and reduced coverage in these regions rather than genuinely low transmission rates.

**Fig. 5:**
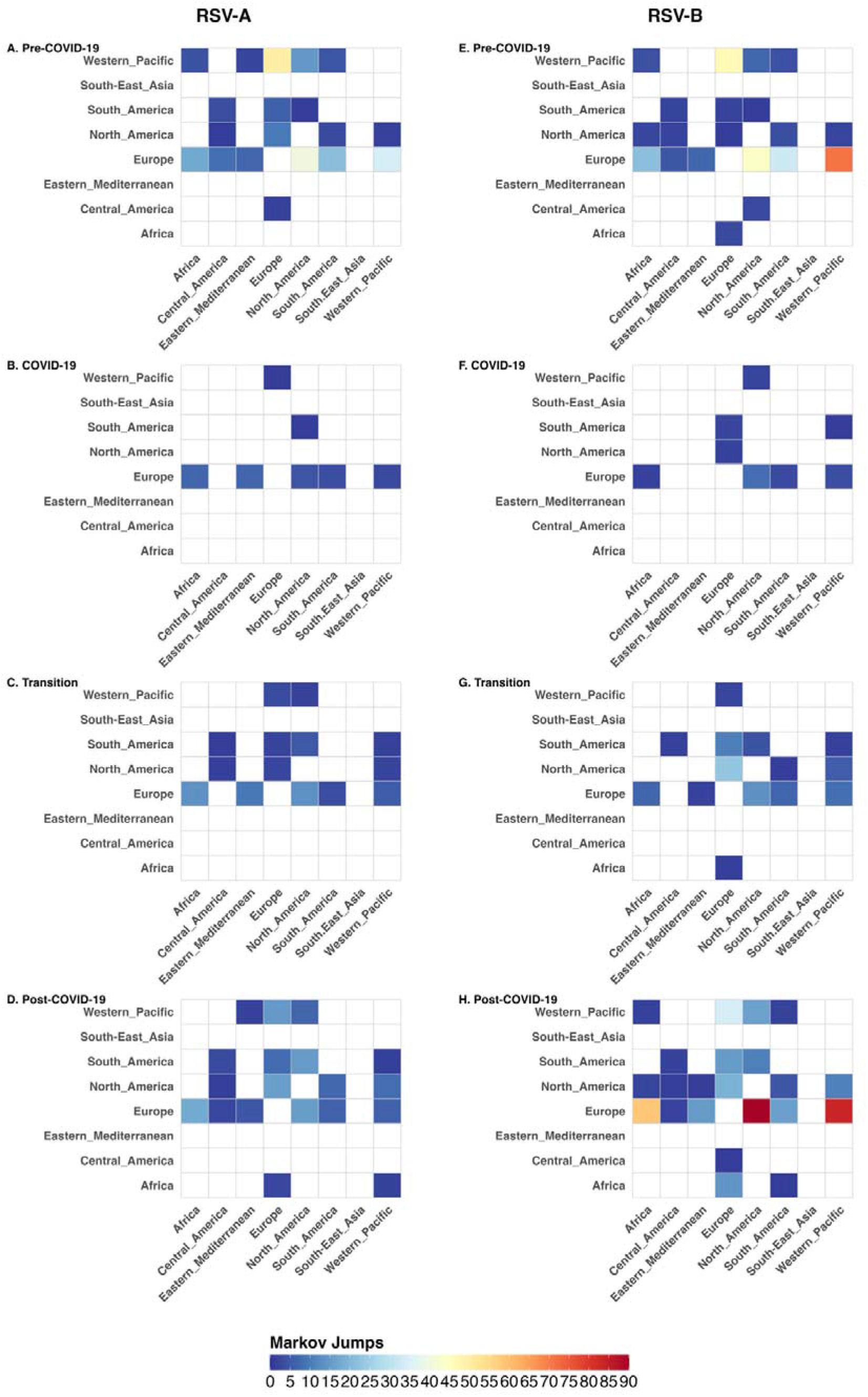
Spatiotemporal migration patterns of RSV-A (A-D) and RSV-B (E-H) across.

RSV-B demonstrated similar pre-pandemic patterns, with Europe (181 events, 95% HPD interval 146-215) and the Western Pacific (65 events, 95% HPD interval 36-86) dominating global transmission. Pandemic suppression was even more pronounced for RSV-B, with European exports reduced to 12 events (95% HPD interval 5-19) and Western Pacific contributions limited to 2-3 introductions in our sample (Fig. 5.E).

During the transition period, transmission dynamics shifted notably. North America emerged as a co-dominant source (30 events, 95% HPD interval 21-40) alongside Europe (38 events, 95% HPD interval 21-56), while South America contributed with 18 events (95% HPD interval 10-27). Post-pandemic recovery showed Europe returning to near pre-pandemic levels (124 events, 95% HPD interval 92-151) (Fig. 5.H), while the Western Pacific (23 events, 95% HPD interval 13-32) and North America (19 events, 95% HPD interval 8-27) maintained comparable export activity to each other.

These findings reveal an uneven global recovery in RSV transmission following pandemic disruptions, reflecting that high-mobility regions, such as Europe, North America and Western Pacific, were resuming their roles as potential sources of international viral dissemination.

### Air travel remains a strong predictor of global RSV circulation

Given unprecedented mobility restrictions during pandemic period, we investigated whether these established relationships persisted across distinct epidemiological contexts. We applied a time-inhomogeneous generalized linear model across four-time intervals or epochs, incorporating air travel volume, population size, and geographic distance as covariates for phylogeographic transitions in each epoch (Fig. 6).

**Fig. 6.**
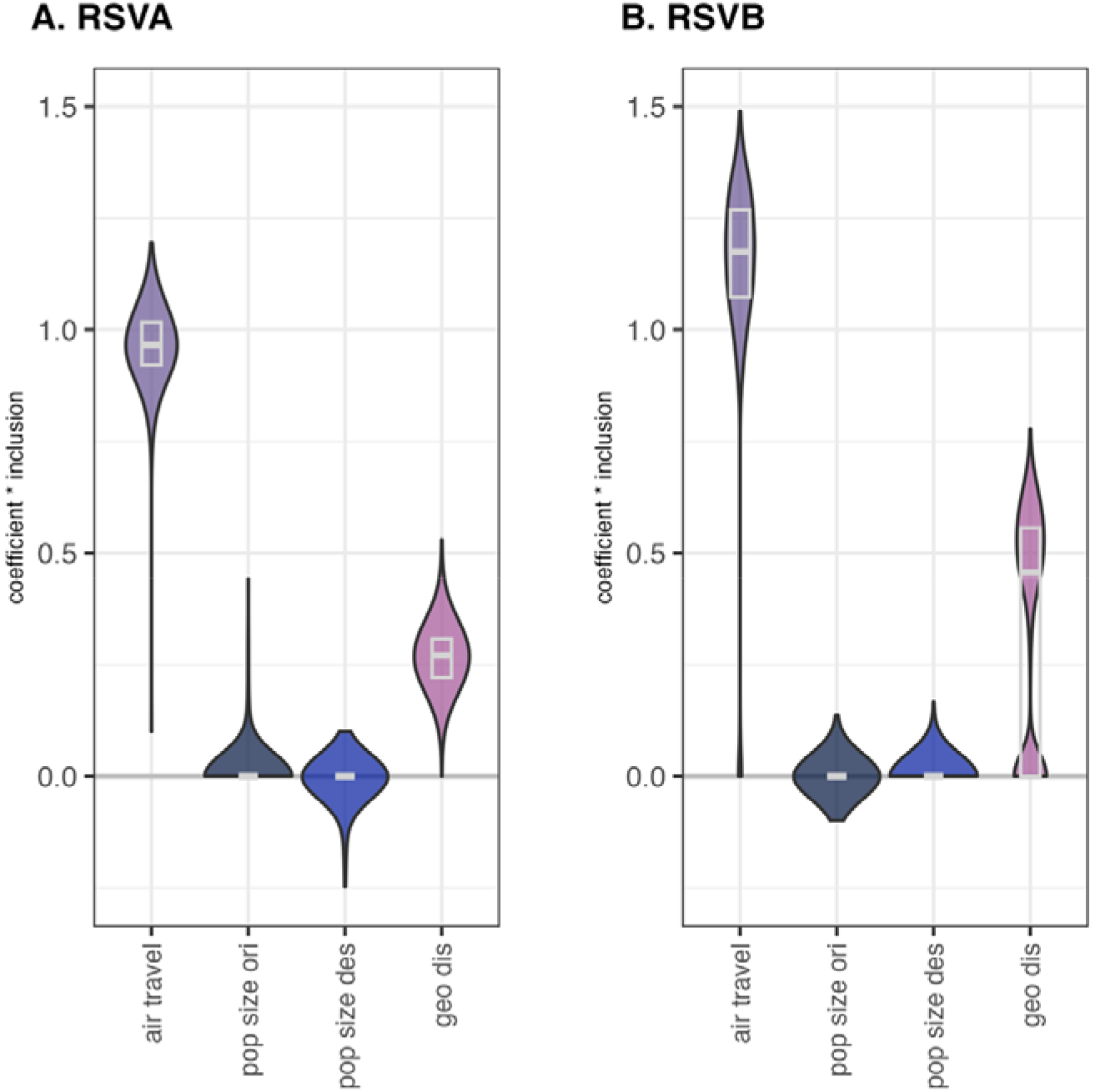
Generalized Linear Model (GLM) analysis of predictors influencing RSV spatial diffusion. Posterior estimates of GLM coefficients for predictors of viral migration rates between geographic regions. Values represent the product of the coefficient (on a log scale) and the inclusion probability for each predictor (coefficient × inclusion). Positive values indicate factors that increase migration rates, while negative values indicate factors that reduce migration rates. The inclusion probability (Bayes Factor) for each predictor is shown on the right. Analysis was performed using an epoch-based GLM framework to account for temporal heterogeneity in migration dynamics across pre-pandemic, pandemic, and post-pandemic periods. Predictors are color-coded as follows: air travel flow (**Slate Purple**), origin population size (**Midnight Navy**), and destination population size (**Royal Blue**) and geographic distance (**Mauve/Purple**).

Air passenger traffic remained a strong predictor of viral dissemination across all periods for both RSV-A (log coefficient 0·97, 95% HPD 0·79–1·10) and RSV-B (log coefficient 0·68, 95% HPD 0·01–0·94). These associations were robust across spatial scales, though population size effects were inconsistent. Geographic distance analysis demonstrated that short geographic distance may also be associated with transmission patterns, particularly during the pandemic period for RSV-A (log coefficient 0·27, 95% HPD 0·12–0.38) and RSV-B (log coefficient 0·33, 95% HPD 0·10–0.64). Our findings confirm that air travel continues to drive global RSV dispersal across lineages and epidemiological contexts, supporting its continued use in predictive surveillance strategies.

Geographic distance analysis yielded an unexpected positive association during the pandemic period for both RSV-A (log coefficient 0·27, 95% HPD 0·12–0·38) and RSV-B (log coefficient 0·33, 95% HPD 0·10–0·64), suggesting more transitions over longer distances.

This counterintuitive result may reflect the disrupted mobility patterns during the pandemic. Nonetheless, our analysis clearly demonstrates that air travel volume was the dominant predictor of viral spread.

### Persistence detection

To assess temporal changes in viral transmission dynamics, we analyzed phylogeographic persistence estimates. Specifically, for four time points of interest during the study period (Table S4), we estimate the proportion of circulating viral lineages that originated from sustained local transmission within a geographic region, relative to those that resulted from new introductions from other regions during the intervening period ^7,8^ (Fig. S1).

For both lineages, we observed elevated persistence levels during both the pandemic and transition periods across most regions, followed by progressive decline through the transition and post-pandemic periods (Fig. 7). Notably, Europe showed the highest values during the pandemic periods and South America demonstrated higher persistence during the transition period compared to the pre-pandemic baseline in both cases (Table S5). In contrast, the post-pandemic period was characterized by the lowest persistence estimates across all regions (Fig. 7).

**Fig. 7.**
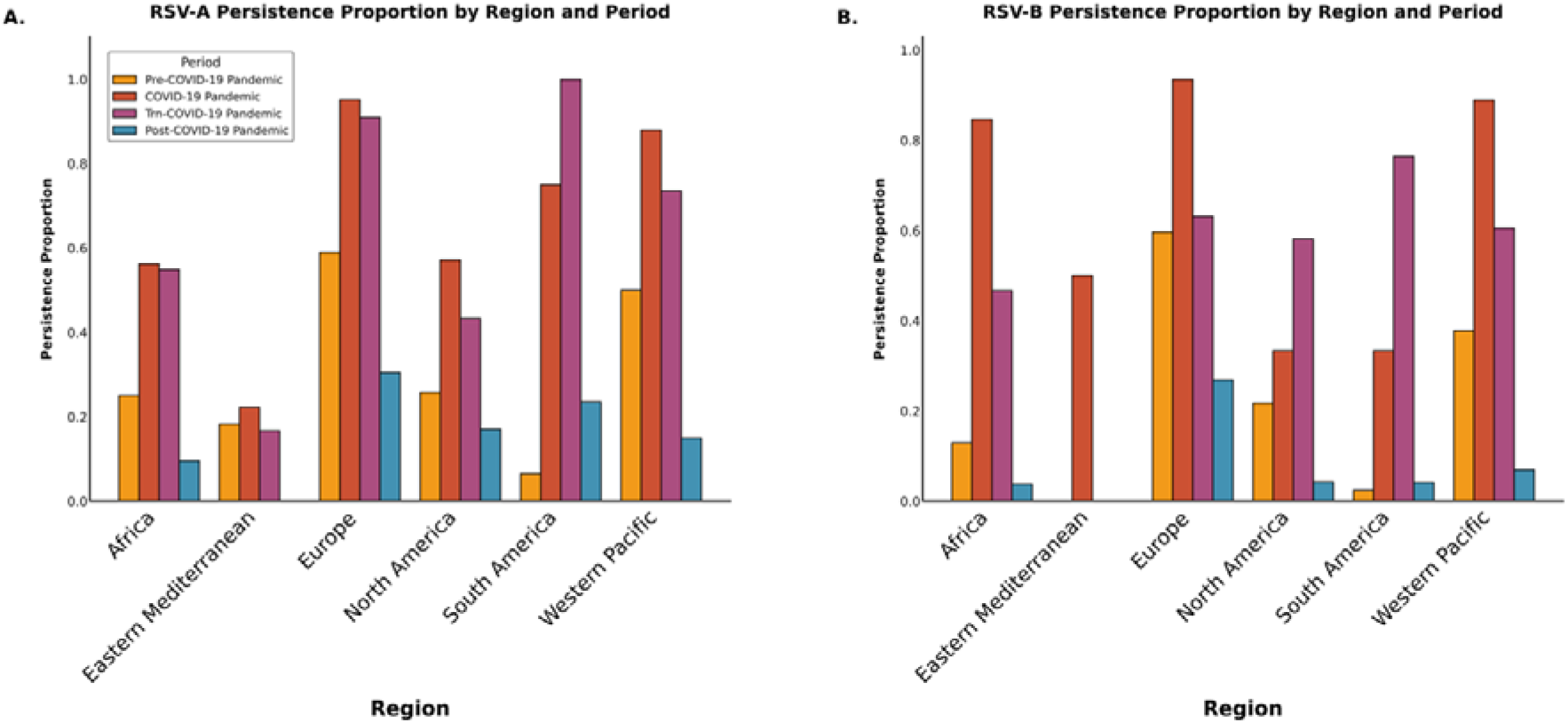
Degree of persistence for RSV-A and B during the Pre, Pandemic, Transition and Post-COVID-19 periods. Stacked bars show proportion of persistence over the periods assessed. Bars are color-based on time periods: Pre-COVID-19 Pandemic (yellow), COVID-19 Pandemic (red), Transition-COVID-19 Pandemic (purple), and post-COVID-19 Pandemic (blue). Central America and Southeast Asia were not included due to insufficient samples.

This pattern suggests that during periods of restricted international travel and stringent NPIs, cross-border viral importation was substantially reduced, allowing locally established transmission chains to predominate and shape the regional genomic diversity of RSV. Conversely, the post-pandemic resurgence was characterized by frequent successful viral introductions that fueled onward transmission, likely driven by resumed international air travel and relaxed public health measures. These findings should be interpreted in the context of limited genomic surveillance data from the African, Eastern Mediterranean and South-East Asian regions, which may affect the generalizability of persistence estimates for these areas.

## Discussion

Our multicenter genomic epidemiology study identified profound disruption of global human RSV transmission dynamics during the pandemic, followed by heterogeneous resurgence patterns for RSV-A and RSV-B, with key shifts in diversity and evolutionary dynamics. Despite substantial reductions in global mobility, air travel remained a crucial driver of international RSV transmission, aligning with patterns observed for other respiratory pathogens including seasonal influenza ^5^. This finding was also supported by the increased persistence we identified during the pandemic and transition periods, indicating that local retention was a relevant source, which is consistent with sustained implementation of NPIs isolating regions, including travel bans in 2020-2021 ^9^. In addition, persistence rates decreased sharply in the post-pandemic phase, indicating an increase in new viral introductions when human mobility recovered ^10,11^.

Both RSV-A and RSV-B demonstrated similar temporal dynamics until mid-2020, when marked population bottlenecks coincided with the onset of the COVID-19 pandemic. RSV circulation only peaked in the following transition period, highlighting a gradual and regionally heterogeneous return of global transmission. While RSV shared mobility-driven transmission patterns with other respiratory viruses, it exhibited distinct geographical source dynamics. European countries consistently served as the predominant sources of global RSV exportation across all study periods (pre-pandemic, pandemic, and post-pandemic) for both viral subtypes, with Western Pacific and North America emerging as secondary sources. Unlike seasonal influenza virus lineage circulation, for which East and Southeast Asia represent a source population ^5^, RSV maintained consistent European dominance regardless of subtype or pandemic phase ^5,10^.

Phylogenetic analysis revealed that specific clades emerged as dominant lineages following the pandemic bottleneck for both RSV subtypes. In the post-pandemic period, clades AD.3 and AD5.2 for RSV-A, and B.D.E.1 for RSV-B expanded, potentially indicating differential fitness advantages. Our site-selection analysis identified distinct evolutionary pressures across these emergent lineages, revealing subtype-specific adaptation strategies with potentially therapeutic implications. While RSV exhibits more conservative evolution compared to the rapid antigenic drift of influenza virus, the pandemic bottleneck accelerated distinct evolutionary trajectories between RSV-A and RSV-B^12^.

Most notably, positive selection at G protein position 273 occurred across multiple RSV-A clades (AD5.2, AD.3, AD3.1), suggesting sustained immune pressure at this critical site. Clade AD.3 showed the most extensive selection with six G protein sites (132, 233, 273, 310, 313, 314) and position 128 in M2-1, while clade AD.1.5 exhibited focused selection at G 284 residue and in the L gene at residue 1524. Positive selection at G protein residue 273 is consistent with recent findings from a whole-genome phylodynamic study of RSV circulation in Maryland, USA (2018–2024), which independently identified this site as being under diversifying selection^13^. This residue also maps to a predicted B-cell epitope region, implicating a functional role in immune recognition. More broadly, the concentration of diversifying selection within the G gene likely reflects sustained pressure from host humoral immunity, whereas the predominance of purifying selection across more conserved genomic regions suggests stringent functional constraints on viral replication and transmission machinery. ^14^.

RSV-B displayed fundamentally different evolutionary dynamics, with the dominant B.D.E.1 clade showing 20 selected sites across multiple proteins. Beyond the expected G protein selection (positions 114, 145, 202, 217, 249, 256, 265, 275, 281). RSV-B exhibited positive selection in regulatory proteins M2-1 (position 175) and M2-2 (position 48), close to the previously identified M2-2 mutations (M2-2:M27T, M2-2: D35N, M2-2:C49F) was identified in RSV-B in the post-pandemic period (here in all B.D.E.1 monophyletic clade with 1618 genomes). The M2-2 protein functions as a regulatory element that modulates host cellular processes to confer selective advantages through enhanced replication efficiency. Through interference with translation initiation mechanisms and interaction with protein degradation pathways, M2-2 optimizes cellular resources for viral synthesis, resulting in increased progeny production and transmission potential. This triggering of key regulatory proteins, including 4E-BP1 and eIF2α, while circumventing host stress responses, establishes conditions conducive to accelerated viral replication ^15^. The potential fitness enhancement may explain the epidemiological persistence of M2-2-variant RSV-B lineages observed in post-pandemic surveillance data. This suggests RSV-B has undergone notable changes in viral fitness architecture that may alter therapeutic susceptibility and warrant intensive post-pandemic surveillance. This dynamic may have been further reinforced by the slightly faster evolutionary rate observed in RSV-B compared to RSV-A ^16^.

Additionally, the population bottleneck induced by pandemic-related interventions may have facilitated a shift within the RSV-B population. In a context of reduced viral competition, lineages with pre-existing fitness advantages may have expanded rapidly, facilitating the emergence and subsequent dominance of RSV-B genotypes.

There are some limitations to be considered when interpreting our findings. Spatial resolution was constrained by genomic data availability and surveillance infrastructure. To mitigate heterogeneous sampling across regions, we employed established global geographical aggregation adapted from WHO and conducted two distinct downsampling strategies (a diversity-based monophyletic approach and an epidemiologically informed approach), both of which supported the reliability of the observed spatial dynamics.

Nevertheless, within-country sample densities were insufficient to characterize fine-scale, short-distance RSV dissemination patterns. Key strengths of this investigation include the leveraging of a unique dataset with robust sample size for RSV-A (n=709 genomes) and RSV-B (n=611 genomes), reliance on complete viral genome sequences, and extensive spatiotemporal coverage spanning multiple years combined with rigorously validated background datasets. Increased sampling density and broader geographic representation will likely reveal additional transmission determinants operating at multiple spatial scales.

## Conclusion

In conclusion, this study presents an integrated genomic-epidemiological framework comparing the pre-and post-pandemic RSV landscape. Our findings identified routes underpinning post-pandemic resurgence, driven by international air travel, and accompanied by marked shifts in viral diversity and evolutionary dynamics. Collectively, these results suggest that the post-pandemic period did not represent a simple reversion to pre-existing period transmission patterns, but rather a profound epidemiological restructuring for RSV-B. These results underscore the relevance of sustained genomic surveillance to monitor emerging variants. A deeper understanding of the evolutionary pressures shaping RSV-B will be essential to optimize current and next-generation interventions globally.

## Author contributions

E.G.-B., H.J.A.A.v.Z., L.B. and P.L. conceived and designed the study. E.G.-B., V.C., A. E., R.J.L., H.J.A.A.v.Z., L.B. and P.L. acquired the data. E.G.-B. and P.L. performed the analyses. E.G.-B., D.M.W., N.D.G., H.J.A.A.v.Z., L.B. and P.L. contributed to data interpretation. E.G.-B., H.J.A.A.v.Z., L.B. and P.L. wrote the original draft. All authors reviewed, edited, and approved the final manuscript.

## Data sharing

A repository with all code used to generate the phylodynamic analyses conducted in this study is available on GitHub: https://github.com/emanuelegustani/Pandemic-Driven-Shifts-in-RSV-Diversity-Evolution-and-Global-Spread.git. Newly generated viral genomes from this study have been deposited under accession numbers RSV-A: XXXXXXXX -XXXXXXXX and RSVB-XXXXXXXX -XXXXXXXX. The background genomic data used in this study will be available in Supplementary File 2.

## Data Availability

A repository with all code used to generate the phylodynamic analyses conducted in this study is available on GitHub: https://github.com/emanuelegustani/Pandemic-Driven-Shifts-in-RSV-Diversity-Evolution-and-Global-Spread.git. Newly generated viral genomes from this study have been deposited under accession numbers RSV-A: QBXXXXX - QBXXXXX and RSVB- QBXXXXX - QBXXXXX. The background genomic data used in this study will be available in Supplementary File 2.

## Acknowledgments

We thank the study participants and their families, along with the research staff from the INFORM-RSV Study Group who contributed to this study. We are especially grateful to Deidre Wilkins and Kelly Mahool for their essential dedication and support throughout the project.

## Methods

### Data Collection

The INFORM-RSV study is a multicenter, global clinical study performed to understand the global epidemiology of RSV in children under 5 years old ^16^with sampling locations across 18 countries. The sampling design comprises 50 or 100 samples per location per year confirmed RSV-positive samples per location per locally defined RSV season using a standardized prospective surveillance protocol, as previously described. The newly sequenced dataset covers the period from mid 2020 to late 2023, comprising 709 RSV-A and 611 RSV-B genomes. The whole genome sequencing was implemented at UMC Utrecht on the Illumina NextSeq500 platform following the protocol previously published ^16^. To extend the temporal scope of the dataset, complementary complete genome sequences obtained between 2017 and early 2020 were retrieved from a previously published INFORM study ^15^.

### Ethical study approval

This study was designed, conducted, and reported in accordance with the principles of the Declaration of Helsinki (2024). Written informed consent was provided by parents or legal guardians before sample collection, in compliance with the International Conference on Harmonisation Guidelines for Good Clinical Practice (ICH-GCP) and all relevant national and international regulatory standards. Ethical approval for the INFORM-RSV study was obtained from the institutional review boards at each of the 18 participating study sites: The Netherlands: The Medical Research Ethics Committee of the UMC Utrecht (reference number WAG/mb/17/016170); Italy: Ethics Committee for Clinical Testing of the Province of Padova of the Padova Hospital (no. 345 of 27/10/2016); Russia: The Department for Science, Innovation Development and Management of Health and Biological Risks, Ministry of Health of the Russian Federation; Germany Marburg (4401): Ethics Committee of the Medical Faculty of the Philipps University Marburg; France Creteil (3301): Ethics Committee

Southwest and Overseas of the Créteil Intercommunal Hospital Centre (ID-RCB No.: 2018-A02360–55 (file 1– 18–73); Spain: Ethics Committee for Research Santiago-Lugo of the Hospital Centre University of Santiago (registration code 2017/397); South Korea: Medical Research Committee of the Seoul National University Hospital; Finland: Ethics Committee of the Hospital District of Southwest Finland, Turku; Australia: Human Research Ethics Committee of the Perth Children’s Hospital; Brazil: The Research Ethics Committee of the Centro INFANT at Pontificia Universidade Catolica de Rio Grande do Sul (opinion number 2,569,872); Canada Hamilton: Hamilton Integrated Research Ethics Board of the McMaster University; Canada: Research Ethics Board of the McGill University Health Centre; Canada Montreal: McGill University Health Centre (MUHC) Research Ethics Board (REB), pediatric panel;Canada Montreal: McGill University Health Centre (MUHC) Research Ethics Board (REB), pediatric panel; South Africa: Human Research Ethics Committee of the University of the Witwatersrand Johannesburg (no. M170966); Japan: Research Ethics Committee of the Fukushima Medical University (no. 29212); United Kingdom (London and Oxford sites): Health Research Authority (HRA) Research Ethics Committee (ref. 17/EM/0469) via the Integrated Research Application System (IRAS), followed by HRA Approval and site-specific Research & Development approvals from King’s College Hospital and Oxford University Hospitals NHS Foundation Trust; Taiwan: Mackay Memorial Hospital Institutional Review Board (no. 19MMHIS171e); Chile: Ethics Committee for Research on Human Subjects of the Faculty of Medicine, University of Chile; Mexico: Ethics Committee of the University Autónoma De Nuevo León, Faculty of Medicine.

### Global Case Surveillance

We analyzed global RSV case data from 2017 to 2024 obtained from the World Health Organization’s FluNet platform (https://www.who.int/tools/flunet). FluNet serves as the global influenza surveillance network, managed by WHO in collaboration with the National Library of Medicine.

### Genomic Background Sequence Dataset

To establish a comprehensive genomic background, we curated a global dataset from GISAID (Supplemental File) comprising 7,081 complete RSV-A and 7,619 complete RSV-B sequences. This dataset spans the period from January 2017 to January 2024, providing a comprehensive genomic context for our analyses. We labeled the following periods: pre-pandemic (2017-2020), pandemic (2020-2021), transition (2021-2022), and relaxation/post-pandemic (2022-2024).

### Lineage and genotype Classification

The lineages were assigned using Nextclade v3.15.3 ^6^, which was also used to assess the genome quality coverage, perform mutation annotation, and to remove poor quality genomes.

### Sequence Alignment and Maximum Likelihood Phylogenetic Reconstruction

Multiple sequence alignment was performed using Nextclade CLI. Maximum likelihood phylogenetic trees were reconstructed using IQ-TREE v2.3.6, with branch support estimated via the SH-aLRT test ^17^ and ultrafast bootstrap approximation ^18^.

### Genomic subsampling

To mitigate spatial sampling heterogeneity and reduce computational burden for Bayesian inference, we applied a dual subsampling strategy to GISAID sequences, while retaining all INFORM study genomes throughout.

The first approach employed PhyloDish ^19^ to identify monophyletic clusters comprising sequences from a single geographic location, randomly downsampling each cluster to a single representative GISAID sequence, yielding datasets of 1,668 sequences for RSV-A and 1,808 for RSV-B. The second approach applied an epidemiological subsampling strategy, selecting sequences proportional to 0.0001% of each location’s population size, sampled at two-day intervals across 2017–2024, with a maximum of 400 sequences per country to prevent geographic overrepresentation, yielding 2,714 sequences for RSV-A and 3,144 for RSV-B. Both strategies yielded concordant phylodynamic patterns, confirming the robustness of our findings. The epidemiological subsampling dataset was carried forward for all downstream analyses.

The final combined datasets, integrating INFORM and GISAID sequences, comprised 3,544 sequences for RSV-A (INFORM: n=709; GISAID: n=2,714) and 3,800 sequences for RSV-B (INFORM: n=611; GISAID: n=3,144), distributed across eight WHO-designated geographic regions as follows: RSV-A: Africa (n=457), Central America (n=48), Eastern Mediterranean (n=65), Europe (n=1,256), North America (n=448), South America (n=336), South-East Asia (n=8), and Western Pacific (n=926). RSV-B: Africa (n=340), Central America (n=31), Eastern Mediterranean (n=41), Europe (n=1,631), North America (n=496), South America (n=252), South-East Asia (n=40), and Western Pacific (n=970).

### Temporal Signal Assessment and Outlier Exclusion

Prior to Bayesian time-measured phylogenetic analyses, molecular clock signal was evaluated in the subsampled datasets using TempEst v1.5.3 ^20^. Outlier sequences deviating substantially from the root-to-tip regression were identified and removed using TreeTime ^21^, resulting in the exclusion of 27 RSV-A and 6 RSV-B sequences. The final root-to-tip correlation coefficients were 0.89 (R²=0.79) for RSV-A and 0.99 (R²=0.98) for RSV-B, confirming strong temporal signal in both datasets and supporting the validity of downstream molecular clock analyses.

### Phylodynamic inference

Time-calibrated phylogenetic reconstruction was performed independently for RSV-A and RSV-B within a Bayesian framework using BEAST X v10.5.0 ^22^, with computational performance enhanced through parallelization via the BEAGLE v4 library ^23^. Identical model specifications were applied to both subtypes. Nucleotide substitution dynamics were modeled using a General Time Reversible (GTR) framework with gamma-distributed among-site rate variation. Among-lineage rate variation was accommodated through an uncorrelated relaxed molecular clock assuming a lognormal distribution of branch rates. Population size dynamics were reconstructed using a Bayesian Skygrid coalescent tree prior, permitting estimation of relative genetic diversity at six-month intervals across the study period. Markov Chain Monte Carlo (MCMC) sampling was conducted over 200 million iterations, with convergence assessed using Tracer v1.7.2 ^24^. Adequate mixing and convergence were confirmed by ensuring effective sample sizes (ESS) exceeding 200 for all parameters.

Maximum clade credibility (MCC) trees were summarized following the removal of an appropriate burn-in fraction, retaining posterior estimates for downstream phylogeographic and lineage persistence analyses.

### Discrete Phylogeographic Reconstruction

To reconstruct the global spatiotemporal dispersal of RSV-A and RSV-B, we implemented a discrete asymmetric phylogeographic model fitted to 1,000 trees randomly drawn from the BEAST posterior distribution for each subtype. Geographic locations were assigned according to WHO regional designations, encompassing eight regions: Africa, Central America, Eastern Mediterranean, Europe, North America, South America, South-East Asia, and Western Pacific, which were incorporated as discrete traits within the phylogeographic model.

Asymmetric phylogeographic reconstruction was performed in BEAST X v10.5.0 ^22^, accelerated via the BEAGLE v4.0.0 library ^23^. Inter-regional dispersal rates were parameterized using an asymmetric Continuous-Time Markov Chain (CTMC) process, with a prior specification on the total number of non-zero rates following the recommendation of Gao and collaborators ^25^. Realized transitions between geographic states were quantified as Markov jump counts. To identify a parsimonious set of well-supported dispersal rates, Bayesian stochastic search variable selection (BSSVS) was applied. MCMC sampling was conducted over 200 million iterations, sampled every 10,000 steps. Convergence was verified in Tracer v1.7.2, with adequate mixing confirmed by ESS values exceeding 200 for all parameters. Posterior logs were combined using LogCombiner v1.10.4 ^22^.

Maximum clade credibility trees were summarized in TreeAnnotator ^22^ and visualized using the ggtree R package ^26^. Markov jump transition histories were extracted from the posterior tree distribution using the TreeMarkovJumpHistoryAnalyzer tool ^11^, restricted to transitions supported by a Bayes factor exceeding 10.

### Uncovering the Drivers of Viral Transmission: Generalized linear mixed model

To assess the factors driving RSV dissemination at the country level, we implemented a GLM extension of discrete phylogeographic diffusion^10^. Our analysis integrates mobility and demographic data to identify key drivers modeling location exchange along a phylogeny as a function of various potential predictors, including air travel, population size, and sample size over specific time points defined in the epoch model (pre-pandemic, pandemic, transition and post-pandemic periods) ^10^. Air mobility data for monthly origin–destination air passenger booking data from 2019 to 2022, were obtained from the International Air Transport Association (IATA). For analysis, volumes were aggregated for relevant time periods per region and country. National population estimates for the 0–4 years age group (both sexes) were obtained from the United Nations World Population Prospects 2024.

### Persistence analysis

Quantifying viral persistence is fundamental for understanding epidemic dynamics and informing public health strategies. Using our phylogeographic reconstructions, we can distinguish between lineages that are the result of an introduction in a particular location and lineages that persisted in that location within a specific time interval. In the former case, a transition on a branch occurred to the location within that time interval, and the descendants of this branch are considered to constitute an introduced lineage. In the latter case, the branches constituting the lineage remained associated with a single location within the time interval (Fig. S1). We summarize the proportion of lineages that persisted over a particular time interval based on the trees annotated with the location transition history from the posterior distribution using PersistenceSummarizer ^11^.

### Identifying site-specific selection

To identify sites evolving under diversifying positive selection, we employed two complementary statistical frameworks implemented in HyPhy v2.5.9.^27^.

The Mixed Effects Model of Evolution (MEME) was applied to detect episodic positive selection operating heterogeneously across branches, permitting selective pressure to vary at the lineage level. The Fast Unconstrained Bayesian Approximation (FUBAR) was employed to identify pervasive diversifying selection acting uniformly across the phylogeny^28^. Significance thresholds were defined as follows: a p-value below 0.1 for MEME, restricting inference to internal branches; a posterior probability exceeding 0.9 for FUBAR. To minimize false positives arising from stochastic episodic events, sites were classified as candidates under positive selection if they were supported by both methods. Only sites meeting the minimum two-method concordance criterion were reported.

### Assessing genealogical neutrality

To assess whether RSV-A and RSV-B lineage distributions during the post-pandemic resurgence period deviate from neutral evolutionary expectations, we implemented a model-based Bayesian posterior predictive framework capable of disentangling the confounding effects of population demographic history from genuine signatures of natural selection.

Posterior tree distributions generated by BEAST were used as the empirical basis for this assessment. Neutral expectation genealogies with same number of tips and sampling time structure were simulated under a Skygrid coalescent model, informed by the empirical parameter estimates, to explicitly account for the complex and temporally heterogeneous demographic trajectories characteristic of RSV populations across pandemic phases ^29^.

To evaluate whether the post-pandemic resurgence lineage dynamics deviated from neutrality, we adopted an entropy metric based on the size distribution of the lineages, with the size being determined by the number tip descendants. Departures from neutral evolution were evaluated by comparing the entropy of the empirically inferred tree distribution against those generated under the neutral posterior predictive model. Statistically significant deviations from the neutral expectation were interpreted as evidence of selection operating the RSV lineages that survived the pandemic bottleneck.

