## Supplemental information 1 for "Pandemic-Driven Shifts in RSV Diversity, Evolution, and Global Spread"

on behalf of the INFORM-RSV Study Group†

†Members listed in the appendix

1.	Canada Montreal

2.	Canada Hamilton

3.	UK London

4.	UK Oxford

5.	Chile

6.	South Korea

7.	Finland

8.	France

9.	Brazil

10.	Mexico

11.	South Africa

12.	Russia

13.	Taiwan

14.	Germany

15.	Italy

16.	Japan

17.	Spain

18.	Australia

19.	Netherlands

Canada Montreal

•	Jesse Papenburg, MD MSc – Montreal Children’s Hospital, McGill University Health Centre, Montréal, Canada

Canada Hamilton

•	Professor Jeffrey M. Pernica, MD – McMaster Children’s Hospital, McMaster University, Hamilton, Canada

•	Jayalakshmi Pothiraj – McMaster Children’s Hospital, Hamilton, Canada

UK London

•	Professor Atul Gupta, MD – King’s College Hospital, London, United Kingdom

UK Oxford

•	Shelley Segal – Oxford University Hospitals NHS Foundation Trust, Oxford, United Kingdom

•	Sarah Hopton – Oxford University Hospitals NHS Foundation Trust, Oxford, United Kingdom

Chile

•	Professor Maria Angélica Palomino, MD – Hospital Roberto del Río, Universidad de Chile, Santiago, Chile

•	Pascale Clement, PhD – Hospital Roberto del Río, Universidad de Chile, Santiago, Chile

Zuid Korea

•	Professor Eun Hwa Choi, MD, PhD – Seoul National University Hospital; Seoul National University College of Medicine, Seoul, South Korea

•	Professor Ki Wook Yun, MD – Seoul National University Hospital; Seoul National University College of Medicine, Seoul, South Korea

Finland

•	Professor Terho Heikkinen, MD, PhD – University of Turku; Turku University Hospital, Turku, Finland

France Creteil

•	Robert Cohen, MD – Université Paris Est Créteil; Centre Hospitalier Intercommunal de Créteil, Créteil, France

•	Corinne Levy, MD – Association Clinique et Thérapeutique Infantile du Val de Marne, Créteil, France

•	Acknowledgement: Professor Naim Ouldali, PhD – Hôpital Robert Debré, Paris, France, Université Paris Cité, Paris, France

Brazil

•	Professor Renato Tetelbom Stein, MD, PhD – Pontifícia Universidade Católica do Rio Grande do Sul, Porto Alegre, Brazil

•	Magali Lumertz, MD – Pontifícia Universidade Católica do Rio Grande do Sul; Hospital Moinhos de Vento, Porto Alegre, Brazil

Mexico

•	Abiel Mascareñas De Los Santos, MD – Universidad Autónoma de Nuevo León; Hospital Universitario Dr. José Eleuterio González, Monterrey, Mexico

•	José Iván Castillo Bejarano, MD – Universidad Autónoma de Nuevo León; Hospital Universitario Dr. José Eleuterio González, Monterrey, Mexico

South Africa

•	Professor Marta Nunes, MD, PhD – South African Medical Research Council; University of the Witwatersrand, Johannesburg, South Africa; Centre International de Recherche en Infectiologie (CIRI), Lyon, France

•	Vicky Baillie, PhD – South African Medical Research Council; University of the Witwatersrand, Johannesburg, South Africa

Russia

•	Daria Danilenko, PhD – Smorodintsev Research Institute of Influenza, Saint Petersburg, Russia

•	Andrey Komissarov, PhD – Smorodintsev Research Institute of Influenza, Saint Petersburg, Russia

Taiwan

•	Professor Hsin Chi, MD, PhD – MacKay Children’s Hospital; MacKay Memorial Hospital Taipei; MacKay Medical University, New Taipei, Taiwan

Germany Marburg

•	Professor Christian Keller, Dr. – University Hospital Giessen and Marburg, Germany; University Hospital Regensburg, Germany (currently working)

•	Monika Bauck, Dr. – University Hospital Giessen and Marburg, Marburg, Germany

Italy:

•	Professor Eugenio Baraldi, MD – University of Padova; Institute of Pediatric Research, Padova, Italy

•	Elena Priante, MD, PhD – University of Padova; Institute of Pediatric Research, Padova, Italy

Japan

•	Koichi Hashimoto, MD, PhD – Fukushima Medical University, Fukushima, Japan

•	Professor Mitsuaki Hosoya, MD, PhD – Fukushima Medical University, Fukushima, Japan

Spain

•	Professor Federico Martinón-Torres, MD, PhD – Hospital Clínico Universitario de Santiago; University of Santiago de Compostela; Instituto de Investigación Sanitaria de Santiago de Compostela; Centro de Investigación Biomédica en Red de Enfermedades Respiratorias; Instituto de Salud Carlos III, Santiago de Compostela/Madrid, Spain

•	Irene Rivero-Calle, MD – Hospital Clínico Universitario de Santiago; Instituto de Investigación Sanitaria de Santiago de Compostela, Santiago de Compostela, Spain

•	Acknowledgement: Santiago study group: Ana Dacosta-Urbieta, Irene Rivero-Calle, Gema Barbeito-Castiñeiras, Alberto Gómez-Carballa, Andres Muy-Pérez, Cintia Alvarez-Smith, Carmen Rodriguez-Tenreiro Sánchez, Lorenzo Redondo-Collazo, Federico Martinón-Torres

Australia:

•	Professor Peter Richmond, MD, PhD – University of Western Australia; The Kids Research Institute Australia; Perth Children’s Hospital; Child and Adolescent Health Service; WA Department of Health, Perth, Australia

•	Ushma Wadia – University of Western Australia; The Kids Research Institute Australia; Perth Children’s Hospital; Fiona Stanley Hospital; WA Department of Health, Perth, Australia

| Supplementary Table 1. Frequency distribution of RSV-A clades identified between 2020–2023 across 17 countries in the INFORM-RSV study. | | |
| --- | --- | --- |
| Clade | **Counts** | **Percentage** |
| A.D.3 | 157 | 22.14 |
| A.D.5.2 | 140 | 19.75 |
| A.D.5.1 | 92 | 12.98 |
| A.D.3.1 | 48 | 6.77 |
| A.D.5 | 46 | 6.49 |
| A.D | 36 | 5.08 |
| A.D.2.1 | 33 | 4.65 |
| A.D.1.8 | 25 | 3.53 |
| A.D.1.5 | 23 | 3.24 |
| A.D.1 | 20 | 2.82 |
| A.D.5.3 | 20 | 2.82 |
| A.D.1.2 | 17 | 2.40 |
| A.D.3.2 | 16 | 2.26 |
| A.D.1.7 | 8 | 1.13 |
| A.D.1.6 | 6 | 0.85 |
| A.D.1.9 | 4 | 0.56 |
| A.D.4.1 | 4 | 0.56 |
| A.D.1.4 | 3 | 0.42 |
| A.D.3.9 | 3 | 0.42 |
| A.D.1.3 | 2 | 0.28 |
| A.D.3.3 | 2 | 0.28 |
| A.D.3.5 | 2 | 0.28 |
| A.D.3.7 | 2 | 0.28 |
| Total | 709 | 100.00 |
| Clades were assigned using the nextclade whole-genome classification system. | | |

| Supplementary Table 2. Frequency distribution of RSV-B clades identified between 2020–2023 across 17 countries in the INFORM-RSV study. | | |
| --- | --- | --- |
| Clade | Counts | Percentage |
| B.D.E.1 | 364 | 59.57 |
| B.D.4.1.1 | 169 | 27.66 |
| B.D.E.4 | 51 | 8.35 |
| B.D.4.1.3 | 18 | 2.95 |
| B.D.E.3 | 5 | 0.82 |
| B.D.E.5 | 2 | 0.33 |
| B.D.E.2 | 1 | 0.16 |
| B.D.E.6 | 1 | 0.16 |
| Total | 611 | 100 |
| Clades were assigned using the nextclade whole-genome classification system. | | |

| **Supplementary Table 3.** Site-specific positive selection analysis of RSV-A clades using FUBAR and MEME methods. Positively selected sites were identified independently using two complementary approaches: FUBAR (Fast Unconstrained Bayesian Approximation), which detects pervasive selection across the entire phylogeny, and MEME (Mixed Effects Model of Evolution), which identifies episodic diversifying selection along individual branches. Sites detected by both methods (Common Sites) represent the highest-confidence candidates for adaptive evolution. Gene-level results are reported for the attachment glycoprotein (G), fusion protein (F), matrix protein (M), transcription anti-termination factor (M2-1), non-structural proteins (NS1, NS2), and the large polymerase (L). Dashes indicate no positively selected sites detected by the respective method. | | | | |
| --- | --- | --- | --- | --- |
| **Clade** | **Gene** | **Common Sites (Both Methods)** | **FUBAR Only** | **MEME Only** |
| **RSV-A** |  |  |  |  |
| **A.D.1.2_clade_i** | G | - | 71 | - |
|  | F | - | 179 | 65 |
|  | M | - | 25 | - |
|  | M2-1 | - | - | - |
| **A.D.1.3_clade_i** | G | **271** | 121 | 281 |
|  | F | - | 121 | - |
|  | NS2 | - | 32 | - |
|  | M | - | 242 | - |
| **A.D.1.5_clade_i** | G | **284** | - | - |
|  | L | **1524** | 1721 | - |
|  | F | - | 119 | - |
|  | M2-1 | - | 182 | - |
|  | NS1 | - | 2 | - |
| **A.D.3.1_clade_i** | G | **273, 284, 297, 298** | 71, 250, 287, 310 | 267 |
|  | SH | **55** | 50 | - |
|  | N | - | 118 | - |
|  | L | - | - | 2032 |
| **A.D.3_clade_ii** | G | **132, 233, 273, 310, 313, 314** | 70, 274, 298, 321 | 142, 298 |
|  | M2-1 | **128** | - | - |
|  | L | - | - | 605 |
| **A.D.5.2** | G | **273, 297** | 106, 225, 235, 260, 295, 314 | 174, 175, 176, 229, 235, 255, 260 |
|  | F | **107** | 325, 406, 553, 562 | 297 |
|  | L | - | - | 1033 |
|  | NS1 | - | - | 55 |
|  | N | - | 216 | 33 |
|  | NS2 | - | 2 | - |
|  | P | - | 231 | - |
|  | M2-2 | - | 74 | - |
| **RSV-B** |  |  |  |  |
| **B.D.E.1** | G | **99, 114, 145, 202, 217, 249, 256, 265, 275, 281** | - | 2, 21, 76, 130, 149, 151, 221, 222, 225, 231, 245, 269, 285, 291, 306 |
|  | F | **42** | 12, 209 | 173, 389 |
|  | N | **216, 243** | - | 65 |
|  | NS1 | - | - | 111, 137 |
|  | NS2 | **11, 26, 48** | - | 118 |
|  | P | - | - | 150, 156 |
|  | M | - | - | - |
|  | SH | **61, 64** | 49 | 15 |
|  | M2-1 | **175** | - | 108, 193 |
|  | M2-2 | **48** | 35 | - |
|  | L | - | 166, 177 | 111, 137 |

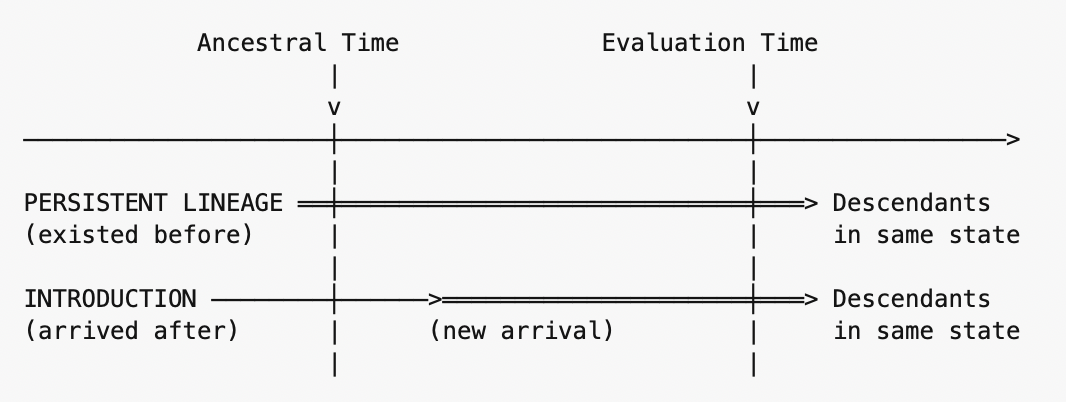

**Figure S1.** Temporal analytical framework for lineage persistence assessment.

Lineages observed at the evaluation time were classified as persistent (originating before the ancestral time; representing continuous local circulation) or introductions (originating between ancestral and evaluation times; representing new importations). The proportion of subsequent local transmission attributable to introduced versus persistent lineages, calculated as the ratio of descendants from introductions to total descendants remaining in the same geographic state.

**Supplementary Table 4.** Summary of periods evaluated.

| Period | evaluationtime | ancestraltime | Calendar window |
| --- | --- | --- | --- |
| Pre-COVID | 4.01 | 4.90 | Feb 2019 → Jan 2020 |
| COVID | 3.01 | 3.90 | Feb 2020 → Jan 2021 |
| Transition | 2.01 | 2.90 | Feb 2021 → Jan 2022 |
| Post-COVID | 0.09 | 1.90 | Feb 2022 → Dec 2023 |

| Supplementary Table S5. Description relative contributions of local persistence versus introduction of RSV circulation in each geographic region | |
| --- | --- |
| Ratio Value | **Interpretation** |
| **High (→1.0)** | Most local transmission after evaluation time came from **new introductions** |
| **Low (→0.0)** | Most local transmission after evaluation time came from **persistent lineages** |
| **~0.5** | Equal contribution from introductions and persistent lineages |
