## Supplemental information 2 for "Pandemic-Driven Shifts in RSV Diversity, Evolution, and Global Spread"

### Supplementary Appendix

All genome sequences and associated metadata supporting the findings of this study can be accessed through the persistent digital object identifier https://doi.org/10.55876/gis8.260324co

In addition to the minted DOI, GISAID also communicates the aggregation of GISAID accession numbers (EPI_ISL_IDs) through the corresponding EPI_SET_260324co identifier to facilitate both, the acknowledgment of all data contributors and the direct retrieval of the underlying data from GISAID used in this study.

#### RSV Data Summary

| GISAID Identifier | Digital Object Identifier | Number of individual viruses | Data Collection range | Number of countries/territories |
|---|---|---|---|---|
| EPI_SET_260324co | https://doi.org/10.55876/gis8.260324co | 4,912 | 2017-01-01 to 2024-01-01 | 51 |
